# From Rule-Based to DeepSeek R1 – A Robust Comparative Evaluation of Fifty Years of Natural Language Processing (NLP) Models To Identify Inflammatory Bowel Disease Cohorts

**DOI:** 10.1101/2025.07.06.25330961

**Authors:** Matthew Stammers, Markus Gwiggner, Reza Nouraei, Cheryl Metcalf, James Batchelor

**Affiliations:** University Hospital Southampton, Southampton, SO16 6YD, United Kingdom; Southampton Emerging Therapies and Technologies (SETT) Centre, Southampton, SO16 6YD, United Kingdom; Clinical Informatics Research Unit (CIRU), Coxford Road, Southampton, SO16 5AF, United Kingdom; University of Southampton, Southampton, SO17 1BJ, United Kingdom, UK; Queen’s Medical Centre, ENT Department, Nottingham, Nottingham, England, NG7 2UH, United Kingdom, UK; University of Southampton, School of Healthcare Enterprise and Innovation, University Of Southampton Science Park, Enterprise Road, Chilworth, SO16 7NS, Southampton, UK

## Abstract

1.1

**Background:** Natural language processing (NLP) can identify cohorts of patients with inflammatory bowel disease (IBD) from free text. However, limited sharing of code, models, and datasets continues to hinder progress, and bias in foundation large language models (LLMs) remains a significant obstacle.

**Objective:** To evaluate 15 open-source NLP models for identifying IBD cohorts, reporting on document-to-patient-level classification, while exploring explainability, generalisability, bias and cost factors.

**Design:** Fifteen algorithms were assessed, covering fifty years of NLP development: regular expressions, Spacy, bag of words (BOW), term frequency inverse document frequency (TF IDF), Word2Vec, two sentence-based SBERT models, three BERT models (distilBERT, RoBERTa, bioclinicalBERT), and five large language models (LLMs): [Mistral-Instruct-0.3-7B, M42-Health/Llama3-8B, Deepseek-R1-Distill-Qwen-32B, Qwen3-32B, and Deepseek-R1-Distill-Llama-70B]. Models were evaluated based on F1 score, bias, environmental costs (in grams of CO2 emitted), and explainability.

**Results:** A total of 9311 labelled documents were evaluated. The fine-tuned DistilBERT model achieved the best performance (F1: 94.06%) and was more efficient (230.1g CO2) than all other BERT and LLM models. BOW was also strong (F1: 93.38%) and very low cost (1.63g CO2). LLMs performed less well (F1: 86.65% to 91.58%) and had a higher compute cost (938.5 to 33884.4g CO2), along with more bias.

**Conclusion:** Older NLP approaches, such as BOW, can outperform modern LLMs in clinical cohort detection when properly trained. While LLMs do not require task-specific pretraining, they are slower, more costly, and less accurate. All models and weights from this study are released as open source to benefit the research community.

## 1.2 Introduction

### 1.2.1 Key Messages

- **What is already known about this topic-** Language models can identify IBD patient cohorts from clinical free-text records, albeit with only moderate accuracy.
- **What this study adds –** This study demonstrates that tried-and-tested NLP methodologies can and do generally outperform LLMs at IBD cohort identification. While they do have higher precision than other methods, LLMs encounter significant speed, bias, and cost limitations. This study also reveals faulty assumptions regarding the simplicity of the document-level free text cohort identification task.
- **How this study might affect research, practice, or policy—**This study highlights significant clinical bias, sustainability, and scalability issues with LLMs, which must be addressed at all levels before practical production clinical deployment can safely occur.

### 1.2.2 Natural Language Processing in Inflammatory Bowel Disease

Ulcerative colitis (UC), Crohn’s disease (CD) and inflammatory bowel disease unclassified (IBD-U) are chronic inflammatory conditions collectively referred to as inflammatory bowel disease (IBD)(1) diagnosed through a combination of clinical, biochemical, genetic, radiological, endoscopic and histopathological tests(2). IBD is known to be less common in non-caucasian populations; however, the incidence of the disease now appears to be increasing(3) among these groups. Natural language processing (NLP) can algorithmically identify patients and perform case matching for clinical trial recruitment(4). However, relatively little robust NLP research has been conducted in gastroenterology to date(5).

Data fragmentation is a known major obstacle(6). Applying NLP algorithms to clinical free text is one of the few ways to address this issue at scale. Rule-based (RB) data trawls using regular expressions (regex) demonstrate high sensitivity but lower precision, with varying overall efficacy across databases (F1: 0.79-0.9). Consequently, better methods are needed to resolve this database fragmentation problem, not only in IBD but also in numerous patient-identification tasks.

Machine learning (ML) NLP algorithms have undergone significant improvements, particularly over the past 50 years. The earliest ML text-classification algorithms took the form of a ‘bag-of-words’ (BOW) word vector representations(7) developed in 1975. These models, in their simplest form, derive counts of words appearing in a document and associate these counts with a class (during training) to later make classification decisions(8). The original pioneers of ML NLP research subsequently devised other methods, such as Term Frequency Inverse Document Frequency (TF-IDF)(9), a form of vector space model(10) focused on rarer words and other similar, simpler NLP document classification models(11).

However, the primary limitation of these classification algorithms was their inability to understand context or complex associations between words. Consequently, despite considerable work between 1990 and 2017, progress in NLP was only incremental. This, however, changed in 2017 with the advent of the transformer architecture(12). The discovery of the attention head finally enabled computers to focus on written context and begin to understand human language better. Within a year of that paper, bidirectional encoder representations from transformers (BERT)(13) and pre-trained generative transformers (GPT)(14) arrived.

DistilBERT(15) is a lighter and faster version of the original BERT model, operating 60% faster while preserving 95% of BERT’s language understanding benchmark performance. In contrast, RoBERTa(16) was trained on over 160GB of uncompressed text. However, while neither of these models was explicitly trained for clinical tasks, BioClinicalBERT(17) was. In contrast, new open-source GPT models are now released weekly and have garnered significantly more public attention since GPT-3(18) and the public release of ChatGPT^TM^ in 2022. Such large language models (LLMs) perform well on closed benchmarks (MedQA, etc.), but their performance on open medical benchmarks is less impressive despite specialist prompting techniques(19).

In this study, we evaluate five of the current open-source frontrunners(20): M42-Health/Llama3-8B(21), Deepseek-R1-Distill-Qwen2.5-32B(22), Qwen3-32B(23), and Deepseek-R1-Distill-Llama3-70B(22) to assess their zero-shot performance against this novel clinical cohort identification task.

### 1.2.3 Aim

This study develops and thoroughly validates open-source document classification models for IBD, exploring the concepts of explainability, cost, and bias in depth.

### 1.2.4 Objectives

1. Develop, validate, and prioritise methods based on RB, ML, and foundation models (LLMs) for identifying IBD patients.
2. Identify biases, economic impacts, and other costs associated with model inference.
3. Investigate interactions in document-level and patient-level cohort identification as well as model explainability.

## 1.3 Methods

### 1.3.1 Inclusion Criteriagas

All adults aged 18 and over who were first electively referred to the tertiary academic teaching hospital for specialist gastroenterology care between 2007 and 2023, and who did not opt out of allowing their clinical data to be used for secondary care research, were considered for inclusion in the study.

### 1.3.2 Checklist and Ethics

The study adheres to the transparent reporting of a multivariable prediction model for individual prognosis or diagnosis (TRIPOD) for AI checklist(24). Full details of this checklist are provided in *Error! Reference source not found.*. The study protocol was submitted to the Wessex REC (23/SC/0152), which provided research ethics board approval. The study was registered locally as RHM MED1947 on 22/03/2023.

### 1.3.3 Primary & Secondary Outcomes

- Primary outcomes of interest were the harmonic F1 score (2 * precision (PPV) * recall (sensitivity) / precision + recall) and Matthews correlation coefficient (MCC) for IBD diagnosis at both patient and document level.
- Secondary outcomes of interest were fairness statistics(DI/EO/DP), time (s), energy (kWh), CO2 production (grams), model Brier scores and Gini coefficients.

### 1.3.4 Study Configuration

#### 1.3.4.1 Data Sources

This study focuses on gastroenterology letters, endoscopy reports, and histopathology reports.

#### 1.3.4.2 Data Pre-Processing and Quality Checking

All data were handled using a SQL pipeline per NHS reproducible analytics pipelines (RAP) best practices(25) using a robust test-driven development (TDD) approach without filtering other than free-text clinical data redaction using Pteredactyl(26), a locally developed Python-based free-text redaction wrapper around Presidio(27), employing the Stanford base de-identifier model(28) to mask patient-identifiable information (PII). The text was then manually screened to remove any PII that Pteredactyl missed(29).

#### 1.3.4.3 Gold Standard Cohort Identification, Data Linkage & Outcome Labelling

A team of three junior doctors, led by a gastroenterology registrar, initially conducted partially blinded manual chart reviews on a randomly selected cohort of suspected IBD patients comprising 2,800 individuals(6). Then, a subset of this cohort, comprising 1,612 patients, was identified, with available and linkable free-text documents.

Free-text documents were chronologically linked, starting with endoscopy records matched to histopathology reports if the procedure occurred within 72 hours before sample receipt, and the histological type aligned. Relevant preceding or following clinic letters were then added. A consultant gastroenterologist (MS, 14 years’ experience) re-validated all linked records, averaging 5.78 documents per patient. A strict IBD definition was applied, with any diagnostic ambiguity (e.g., ‘possible’ or ‘potential’ IBD) deemed non-diagnostic to maximise classifier precision. Because the primary purpose of the algorithm was to identify all patients who should be under our local IBD service, patients with microscopic colitis were included in the capture, but all other forms of colitis were excluded. Microscopic colitis is strongly associated with IBD(30). At an IBD service planning level, it is appropriately included. However, there may be a demand for these models to be used at a population level in future. Re-training the models to achieve this would be a relatively trivial matter.

#### 1.3.4.4 Predictor Handling

The training and validation sets for the trained models were divided 70/30 (at a patient level), and each model’s 30% holdout set was utilised exclusively for validation with checks in place to prevent data leakage. However, LLMs were evaluated against the entire set because this study for them represents type IV validation(31) because there is no way these LLMs could have ever seen the source data before.

#### 1.3.4.5 Platform Hardware, Software & Dev-Ops

The platform itself was set up as described in *Error! Reference source not found.*. Here, you can find all the links to the code, along with detailed information about the physical hardware setup.

#### 1.3.4.6 LLM Prompt Templating

A JSON-based zero-shot query method was employed to assess the LLMs, and this process is described in more detail in *Error! Reference source not found.*. This template enabled attempts to be made to assess LLM calibration, and by building on MedPrompt(19), leveraging the Clue and Reasoning Prompt (CARP) method(32), facilitating state-of-the-art document classification performance.

### 1.3.5 Analytical Methods

#### 1.3.5.1 Sample Size Calculation

The outcome of this study is a binary classification between ‘IBD’ and ‘Not-IBD’. The most significant factors influencing sample size are class imbalances in the training and validation cohorts, which were nearly 8 to 2 IBD to Not-IBD(6).

Using Pate & Riley’s formula(33) with a predicted r^2^ of 0.05, shrinkage of 0.9, a prevalence of 0.8 and 4 candidate predictors (4 data streams with clinic letters preceding and following the endoscopies/histopath are analysed separately), a sample size of 542 is suggested as required to power the study sufficiently. However, the formula above likely underestimates the actual number of candidate predictors, which cannot be known at this point. Juckett et al.’s (34) work suggests that rare tokens carry less predictive weight and that once samples exceed 1,000 records, a capture probability of> 95% is typically attained. If all 4 document types were vastly different, then a minimum sample size of 4,000 would be required. Because a consultant(attending) physician was performing the study, bringing down the cost of labelling, in the end, nearly 10,000 documents were annotated to absolutely guarantee sufficient power for the study.

#### 1.3.5.2 Study Metrics

A complete set of measurement metrics is used in this study, as highlighted in *Error! Reference source not found.*.

#### 1.3.5.3 Statistical Analyses

Means and medians were calculated as appropriate based on skewness, using a 95% confidence interval or 25%/75% quantiles. Model performance was compared by age, sex, and the index of multiple deprivations decile (IMD), with 10 being the least deprived. Statistical tests were performed as appropriate, with Chi2, Pearson correlation, and Mann-Whitney U tests being used predominantly due to the non-parametric nature of the data. ‘Wilson’s confidence intervals for binomial proportions were calculated using the stats module from Scipy, which provides better coverage probabilities than a normal approximation.

#### 1.3.5.4 Document/Patient-Level Interactions

Decision tree (DT) algorithms were used to determine the optimal fit between document-level and patient-level predictions. Gini coefficients assessed the purity of each branch in the tree’s logic, with a value of 0, indicating perfect separation. Logistic Regression (LR) classifiers were used in many of the NLP pipelines, but these are described in more detail below in ***Model Setup***.

#### 1.3.5.5 Handling Class Imbalance

Due to class imbalance, the harmonic F1 score was preferred as the primary metric for outcome measurement, followed by the Matthews Correlation Coefficient (MCC).

#### 1.3.5.6 Cross Validation & Calibration

Discrimination was assessed using the Brier score(35) and model calibration was evaluated visually using calibration plots. Cross-validation was performed as per ***Predictor Handling.*** Feature selection was performed by the models as described in ***Model Setup***, with the final designs decided upon after much experimentation and records of the older experiments retained.

#### 1.3.5.7 Missingness Transformations & Protecting Clinical Free-Text Information

The text was fully pseudonymised as described in ***Data Pre-Processing and Quality Checking***. Protecting PII was prioritised over any negative impacts on algorithm performance caused by masking. Missingness was quantified and emphasised where relevant. No other transformations were applied to the data as it did not require winsorising or scaling.

#### 1.3.5.8 Error Analysis

Any significant outliers or abnormal results were re-examined through carefully stored outputs in folders throughout the pipeline, along with tests to manage the system. The code was fully documented with docstrings and complete tests to minimise errors. If an issue is discovered, please raise it as an issue on the <u>GitHub page.</u>

#### 1.3.5.9 Fairness/Bias Evaluation

Fairness evaluation was conducted on binned demographic characteristics for every model using demographic parity (DP)(36), equal opportunity (EO)(37) and disparate impact (DI)(38) statistics. All fairness analyses were performed on the validation set only.

#### 1.3.5.10 Economic & Sustainability Analysis

Inference time and computation costs were calculated for each model in succession(39). Emission factors are derived from UK government statistics and the 2024 conversion factors(40), which are set at 0.20705 kg CO2e per kWh according to the 2024 guidance to provide a level playing field for comparison. Calculating the precise energy usage and carbon footprint of LLMs is more challenging because they typically operate across multiple GPUs simultaneously, with tests often taking several days to complete through a local network API, which can be more easily interrupted. Therefore, the best estimates were derived using average watt consumption per hour and algorithm runtime.

#### 1.3.5.11 Explainability Analysis

To better understand model predictions, SHapely Additive exPlanations (SHAP)(41,42)-2014 and Local Interpretable Model-agnostic Explanations(LIME)(43)-2016 were both used. These are the two most popular ML explainability methods presently available(44).

### 1.3.6 Model Setup

Fifteen models are analysed in this study, falling into three primary groups. The first five models [regex, spacy, bag-of-words (BOW)(7), term frequency-inverse document frequency (TF-IDF)(9) and word-2-vector (word2vec)] models are all trained from scratch. The next five transformer-based models are all fine-tuned: [sBERT(45), sBERT-med(46), distilBERT(15), bio-clinicalBERT(15)[BioClinicalBERT] and RoBERTa(47)] and the final five, all GPT-based models were managed solely via prompt engineering [Mistral-Instruct-0.3-7B(20), M42-Health/Llama3-8B(21), Deepseek-R1-Distill-Qwen2.5-32B(22), Qwen3-32B(23), and Deepseek-R1-Distill-Llama3-70B(22)].

Part of the reason for publishing the code fully open source is to allow other developers and data scientists to inspect for themselves how each algorithm was handled. However, for convenience, a complete description of the handling of each model is provided in *Error! Reference source not found.* to make this explicit for all readers.

## 1.4 Results

### 1.4.1 Total Study Cohort

From the randomly selected gold-standard cohort of 2,800 patients, 1,612 individual patients were found to have chronologically linkable endoscopic and histopathological records, as per ***Figure 1***. Eighty-nine patients did not have chronologically linkable clinic letters available.

**Figure 1:**
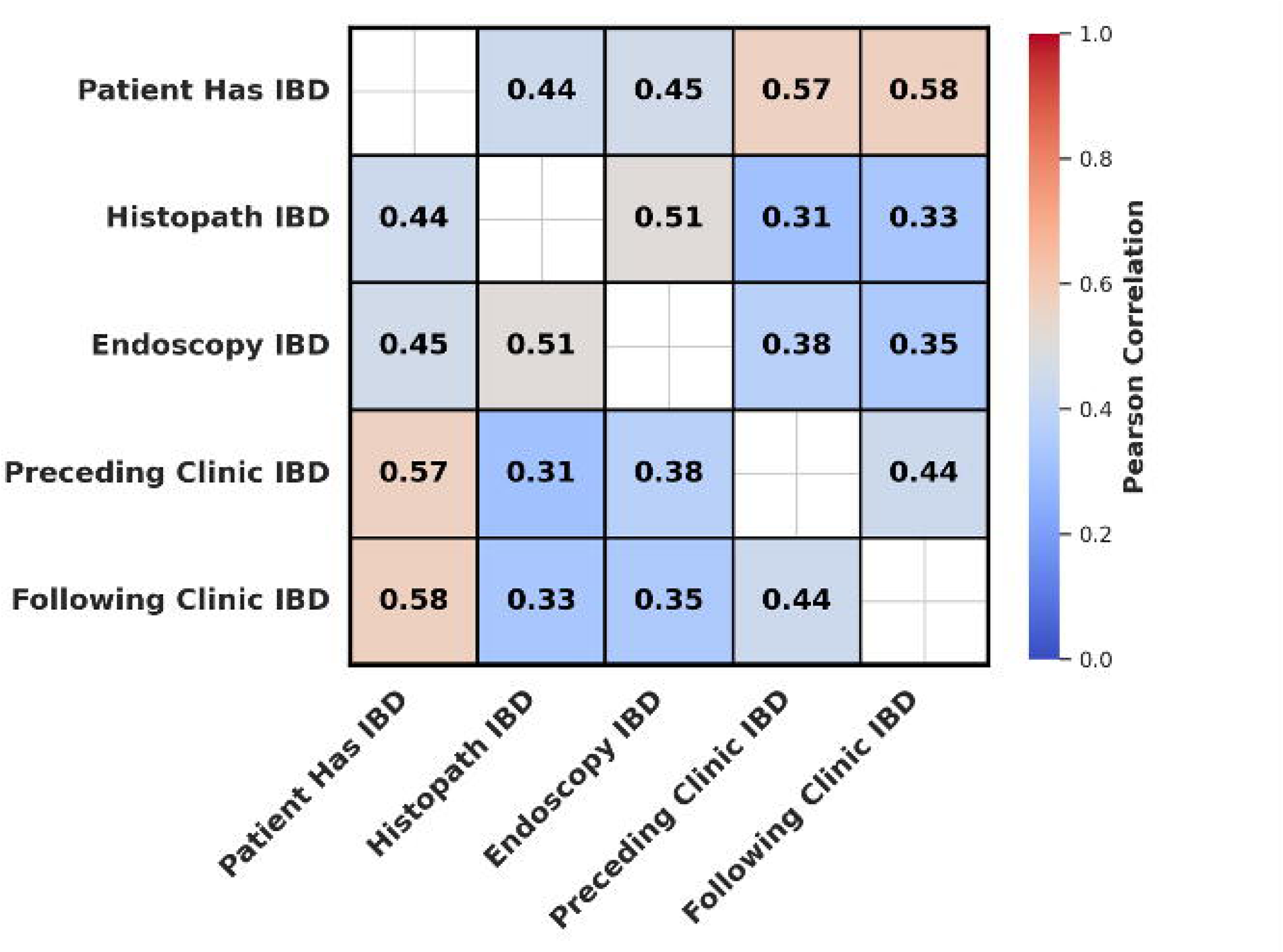
Study Population CONSORT. Figure 1 describes the study population in a CONSORT diagram

#### 1.4.1.1 Training and Validation Cohorts

The training set contained 1,128(70%) patients, and the validation set contained 484(30%). Within the training set, 872(77%) of patients had IBD, and 258(23%) did not. Within the validation set, 378(78%) of patients had IBD, and 110(22%) did not.

In total, 9,311 free-text documents were manually reviewed. The training set contained 6,559 documents, of which 4492(68%) were labelled as suggestive of IBD and 2097(32%) were not. The validation set contained 2,752 free-text documents, of which 1796(65%) were labelled as suggestive of IBD and 956(35%) were not. There were 2,592 rows of carefully aligned document data – 1824 in the training set and 768 in the validation(test) set. Coverage is reported according to the number of these rows used. There were no significant differences between the development and validation settings, eligibility criteria, outcome and predictors.

#### 1.4.1.2 Cohort Demographics

No significant differences were observed between the training and validation cohorts in any continuous demographic. However, the validation cohort had ∼3% more females, and the validation cohort was ∼2% less white. These were the only two significant results. Full demographic results are provided in *Error! Reference source not found.*.

### 1.4.2 Predictive modelling

#### 1.4.2.1 UMLS vs Free Text

The addition of UMLS had a detrimental effect on overall performance. This is because the meta-thesaurus, even though it was carefully filtered, still captured some terms inappropriately connected to IBD, such as ‘17-hydroxycorticosterone’ and ‘vinblastine/methotrexate’, which all map to IBD-associated concept unique identifiers (CUIs) within UMLS. The PTs are thus vulnerable to over-mapping across ontologies. Accordingly, only the free-text results are reported in full here.

#### 1.4.2.2 LLM Response Quality by Model

m42 provided the most incorrectly formatted .json responses n=391, followed by mistral, n=5. The larger LLMs rarely produced incorrectly formatted .json responses (n=2).

#### 1.4.2.3 Document Level Full Results

At a document level, model performance was very variable. Top performers included BOW (F1: 93.84%), SBERT (F1: 94.25%) and the full BERT models (F1: 94.47-95.13%). Full results are given in *Error! Reference source not found.*. Most models lack specificity, indicating that they are not very effective at correctly identifying true negatives.

Full development and validation results for each model are provided in the GitHub repo for complete transparency.

#### 1.4.2.4 Patient-Level Full Results With Patient-Level Trained Models

At a patient level, model performance degrades by approximately 2% of the F1 score if the models are not retrained and 1% if they are (***Table 1***). BOW and the ‘all-mpnet-base-v2’ SBERT model performed exceptionally well. Interestingly, the medically finetuned SBERT model performed comparatively poorly and RoBERTa suffered a catastrophic collapse during the patient-level prediction task.

**Table 1:** Patient Level Prediction Results.

| Model | Doc Coverage | Accuracy | Precision | Recall | Specificity | NPV | F1 Score | MCC |
| --- | --- | --- | --- | --- | --- | --- | --- | --- |
| **Fully Trained** |  |  |  |  |  |  |  |  |
| **Regex** | 768<br>(100.00%) | 81.51%<br>(CI:<br>78.61% -<br>84.10%) | 81.24%<br>(CI:<br>78.31% -<br>83.86%) | 100.00%<br>(CI:<br>99.38% -<br>100.00%) | 7.19% (CI:<br>4.06% -<br>12.41%) | 100.00%<br>(CI:<br>74.12% -<br>100.00%) | 89.65%<br>(CI:<br>89.26% -<br>90.04%) | 0.2417<br>(CI:<br>0.1623 -<br>0.3016) |
| **Spacy** | 768<br>(100.00%) | 80.99%<br>(CI:<br>77.26% -<br>84.24%) | 80.34%<br>(CI:<br>76.50% -<br>83.69%) | 100.00%<br>(CI:<br>98.99% -<br>100.00%) | 14.81% (CI:<br>9.33% -<br>22.72%) | 100.00%<br>(CI:<br>80.64% -<br>100.00%) | 89.10%<br>(CI:<br>88.37% -<br>89.84%) | 0.3450<br>(CI:<br>0.2568 -<br>0.4168) |
| **BOW** | 768<br>(100.00%) | 88.84%<br>(CI:<br>85.73% -<br>91.35%) | 89.02%<br>(CI:<br>85.70% -<br>91.64%) | 98.20%<br>(CI:<br>96.32% -<br>99.12%) | 51.04% (CI:<br>41.20% -<br>60.81%) | 87.50%<br>(CI:<br>76.37% -<br>93.81%) | 93.38%<br>(CI:<br>92.16% -<br>94.59%) | 0.6138<br>(CI:<br>0.5201 -<br>0.7015) |
| **TF-IDF** | 768<br>(100.00%) | 83.68%<br>(CI:<br>80.12% -<br>86.70%) | 83.37%<br>(CI:<br>79.71% -<br>86.48%) | 99.48%<br>(CI:<br>98.14% -<br>99.86%) | 19.79% (CI:<br>13.05% -<br>28.86%) | 90.48%<br>(CI:<br>71.09% -<br>97.35%) | 90.72%<br>(CI:<br>89.86% -<br>91.69%) | 0.3773<br>(CI:<br>0.2729 -<br>0.4738) |
| **Word2Vec** | 768<br>(100.00%) | 81.82%<br>(CI:<br>78.14% -<br>85.00%) | 81.51%<br>(CI:<br>77.78% -<br>84.74%) | 100.00%<br>(CI:<br>99.02% -<br>100.00%) | 8.33% (CI:<br>4.28% -<br>15.59%) | 100.00%<br>(CI:<br>67.56% -<br>100.00%) | 89.81%<br>(CI:<br>89.30% -<br>90.44%) | 0.2606<br>(CI:<br>0.1588 -<br>0.3340) |
| **Finetuned** |  |  |  |  |  |  |  |  |
| **SBERT** | 768<br>(100.00%) | 89.67%<br>(CI:<br>86.64% -<br>92.08%) | 88.81%<br>(CI:<br>85.44% -<br>91.48%) | 99.20%<br>(CI:<br>97.68% -<br>99.73%) | 56.48% (CI:<br>47.07% -<br>65.45%) | 95.31%<br>(CI:<br>87.10% -<br>98.39%) | 93.72%<br>(CI:<br>92.50% -<br>94.90%) | 0.6844<br>(CI:<br>0.6120 -<br>0.7545) |
| **SBERT Med** | 768<br>(100.00%) | 84.71%<br>(CI:<br>81.23% -<br>87.64%) | 84.58%<br>(CI:<br>80.97% -<br>87.61%) | 98.97%<br>(CI:<br>97.38% -<br>99.60%) | 27.08% (CI:<br>19.20% -<br>36.73%) | 86.67%<br>(CI:<br>70.32% -<br>94.69%) | 91.21%<br>(CI:<br>90.07% -<br>92.38%) | 0.4308<br>(CI:<br>0.3263 -<br>0.5233) |
| **DistilBERT** | 768<br>(100.00%) | 90.08%<br>(CI:<br>87.10% -<br>92.44%) | 90.48%<br>(CI:<br>87.29% -<br>92.93%) | 97.94%<br>(CI:<br>95.98% -<br>98.95%) | 58.33% (CI:<br>48.34% -<br>67.69%) | 87.50%<br>(CI:<br>77.23% -<br>93.53%) | 94.06%<br>(CI:<br>92.59% -<br>95.30%) | 0.6624<br>(CI:<br>0.5724 -<br>0.7458) |
| **BioclinicalBERT** | 768<br>(100.00%) | 90.29%<br>(CI:<br>87.33% -<br>92.62%) | 91.48%<br>(CI:<br>88.39% -<br>93.81%) | 96.91%<br>(CI:<br>94.67% -<br>98.22%) | 63.54% (CI:<br>53.57% -<br>72.48%) | 83.56%<br>(CI:<br>73.43% -<br>90.34%) | 94.12%<br>(CI:<br>92.79% -<br>95.48%) | 0.6735<br>(CI:<br>0.5892 -<br>0.7538) |
| **RoBERTa** | 768<br>(100.00%) | 80.17%<br>(CI:<br>76.38% -<br>83.47%) | 80.17%<br>(CI:<br>76.38% -<br>83.47%) | 100.00%<br>(CI:<br>99.02% -<br>100.00%) | 0.00% (CI:<br>0.00% -<br>3.85%) | nan%<br>(CI: N/A) | 88.99%<br>(CI:<br>88.99% -<br>88.99%) | 0.0000<br>(CI:<br>0.0000 -<br>0.0000) |

Table 1 Describes the full patient-level analysis results for the various NLP models in the study.
| Prompt Engineered |  |  |  |  |  |  |  |  |
| --- | --- | --- | --- | --- | --- | --- | --- | --- |
| **Mistral-0.3-7B** | 2510<br>(100.00%) | 81.05%<br>(CI:<br>79.48% - 82.53%) | 94.47%<br>(CI:<br>93.30% - 95.44%) | 81.37%<br>(CI:<br>79.63% - 83.00%) | 79.67% (CI:<br>75.85% - 83.02%) | 50.07%<br>(CI:<br>46.54% - 53.59%) | 87.43%<br>(CI:<br>86.34% - 88.56%) | 0.5214<br>(CI:<br>0.4850 - 0.5571) |
| **M42-Llama_8B** | 2510<br>(100.00%) | 81.67%<br>(CI:<br>80.11% - 83.14%) | 93.26%<br>(CI:<br>92.01% - 94.33%) | 83.32%<br>(CI:<br>81.63% - 84.88%) | 74.79% (CI:<br>70.74% - 78.46%) | 51.71%<br>(CI:<br>48.01% - 55.40%) | 88.01%<br>(CI:<br>86.91% - 89.10%) | 0.5112<br>(CI:<br>0.4755 - 0.5486) |
| **DeepSee k-R1-Qwen2.5_32B** | 2510<br>(100.00%) | 80.31%<br>(CI:<br>78.72% - 81.80%) | 95.83%<br>(CI:<br>94.78% - 96.68%) | 79.08%<br>(CI:<br>77.27% - 80.78%) | 85.48% (CI:<br>82.08% - 88.33%) | 49.18%<br>(CI:<br>45.83% - 52.53%) | 86.65%<br>(CI:<br>85.54% - 87.73%) | 0.5391<br>(CI:<br>0.5084 - 0.5672) |
| **Qwen3_32B** | 2510<br>(100.00%) | 80.22%<br>(CI:<br>78.63% - 81.72%) | 96.35%<br>(CI:<br>95.34% - 97.15%) | 78.50%<br>(CI:<br>76.67% - 80.22%) | 87.47% (CI:<br>84.24% - 90.12%) | 49.13%<br>(CI:<br>45.82% - 52.46%) | 86.51%<br>(CI:<br>85.27% - 87.67%) | 0.5478<br>(CI:<br>0.5178 - 0.5774) |
| **DeepSee k-R1-Llama70B** | 1326<br>(52.82%) | 86.95%<br>(CI:<br>84.87% - 88.77%) | 96.68%<br>(CI:<br>95.25% - 97.69%) | 86.99%<br>(CI:<br>84.69% - 89.00%) | 86.73% (CI:<br>81.49% - 90.66%) | 60.00%<br>(CI:<br>54.41% - 65.34%) | 91.58%<br>(CI:<br>90.25% - 92.89%) | 0.6464<br>(CI:<br>0.5979 - 0.6938) |

LLMs were generally better at patient-level prediction but struggled with document-level prediction vs other models, suggesting that they perform better with greater available context.

#### 1.4.2.5 Model Calibration

Brier scores ranged from 0.711 (DistilBERT) to 0.121 (SBERT Med) for the document classification task and from 0.093 (DistilBERT) to 0.1983 (RoBERTa) on the patient-level prediction task. RoBERTa was trained identically to the other two BERT models with the same hyperparameters. Its catastrophic collapse is purely due to differential pre-training and tokenisation, leading it to predict only positives correctly at a patient level. Such is the potentially brittle nature of these algorithms. Clinical pre-training did not cause the bioclinicalBERT model to outperform distilBERT. The same was true with the SBERT models, where the base SBERT algorithm was substantially better calibrated (0.0829-0.1178) than the medically pre-trained one (0.1061-0.1954). BOW also performed remarkably well (0.098-0.124) vs other ML algorithms like word2vec (0.1211-0.1901).

LLMs, despite not having been fine-tuned to the task, performed remarkably well. However, their Brier scores ranged from 0.1305 (Deepseek Llama 70B) to 0.1978 (Deepseek Qwen 32B), suggesting that their predictions were comparatively poorly calibrated. This will be assessed in more detail in a subsequent study.

### 1.4.3 Document and Patient-Level Interactions

Notably, performance in the document classification task does not reliably predict performance at the patient-level identification task.

#### 1.4.3.1 Document and Patient-Level Database Correlations

Correlations between individual document types and diagnosis vary dramatically even by database. These effects are highlighted in ***Figure 2***, with clinic letters more strongly correlated (0.57-58) with patients ultimately having IBD than endoscopy reports (0.45) or histopathology reports (0.44).

**Figure 2:**
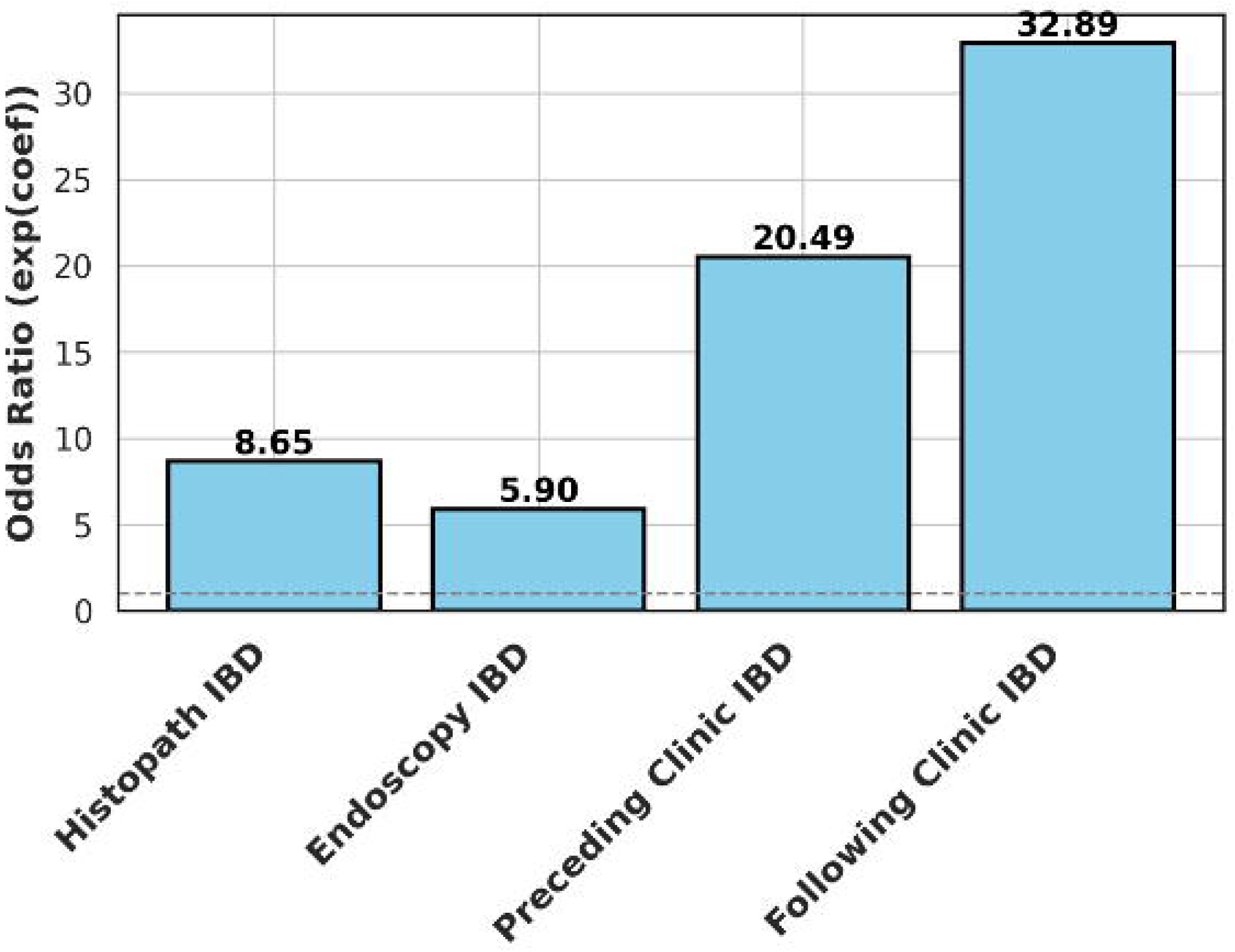
Pearson Correlations Between Document and Patient-Level IBD Diagnosis. Figure 2 describes the Pearson correlations between gold-standard cohort elements

This complex, unexpected complication has not even been mentioned in the literature up until this point(5,6,48). These complexities explain a lot of the performance degradation seen when moving from document to patient-level prediction.

#### 1.4.3.2 Document to Patient Regression and Tree Models

A simple L1 (lasso) LR model was used to assess the individual comparative predictive performance of each variable towards a correct diagnosis of IBD. The following clinic letter was the most useful, with odds of 32.89 vs 20.49 for the preceding letters and 5.9-8.65 for endoscopy and histopathology records (***Figure 3***).

**Figure 3:**
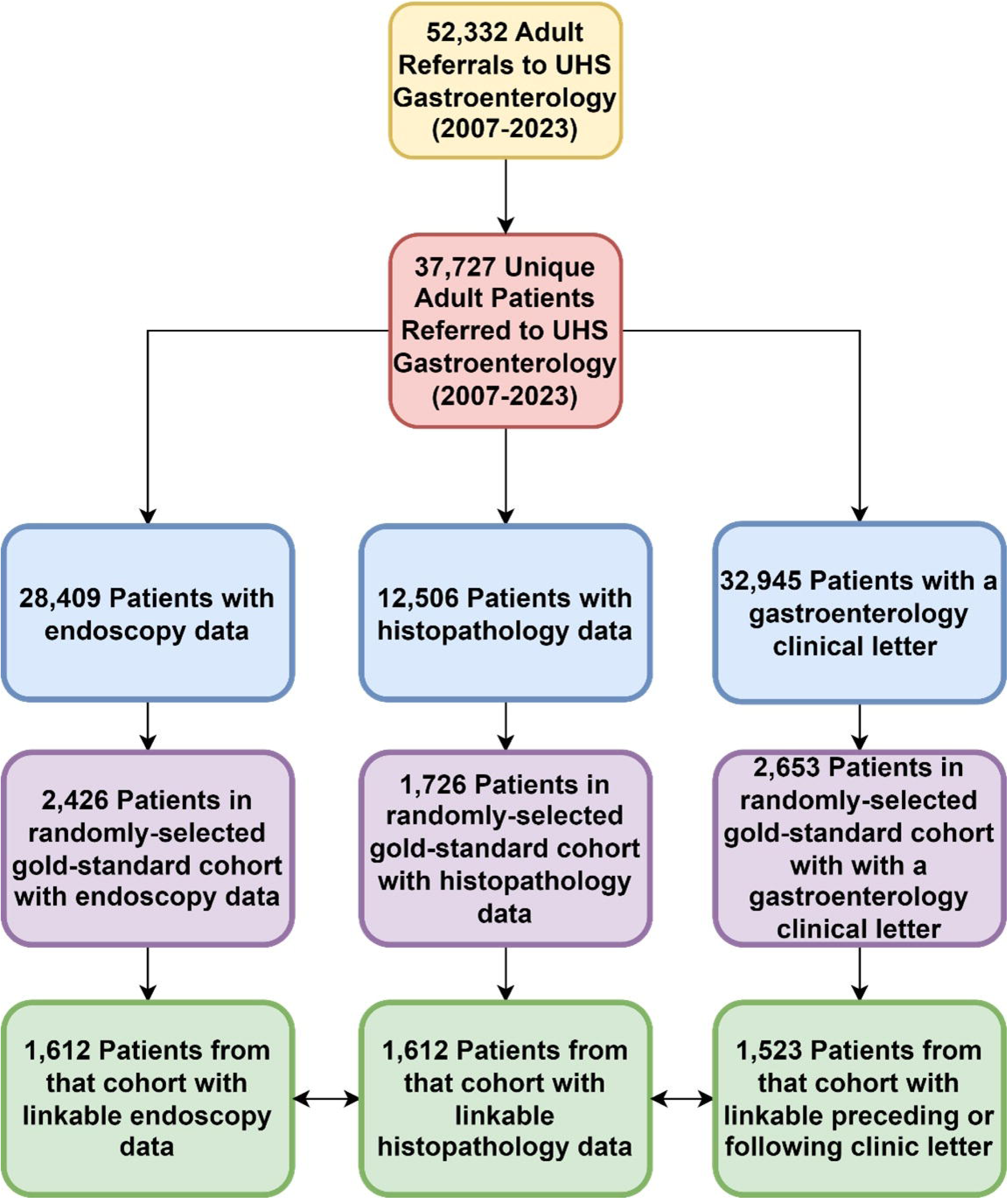
Odds Ratios by Document Type. Figure 3 above highlights the different weights of positive document identification as a contributor to patient-level IBD prediction.

Finally, a decision tree classifier was developed to visualise and attempt to manage the above matrix if possible. However, the gini coefficient never reached zero at any branching step of the logic tree, even if the following clinic split represented the first branch, suggesting that document-to-patient level mapping is not simply solved.

#### 1.4.3.3 Document to Patient Model Performance Variation

Clinic letters appeared to be the most difficult to classify Preceding: (MCC: 0.279-0.678), following: (MCC:0.173-0.779) with endoscopy being the easiest (MCC: 0.611-0.776) and histopathology being only slightly more difficult (MCC: 0.543-0.763). This suggests that the document models tend to perform best for document types that are less important in predicting whether individual patients have IBD overall. Although individual models excelled at specific document types, only distilBERT and bioclinicalBERT sustained MCCs above 0.6 across the options presented, suggesting a significant degree of brittleness among the models tested in this experiment.

### 1.4.4 Fairness Analysis

In *Error! Reference source not found.* the full fairness results are given for each model at a patient level. Overall, LLMs were the most biased, followed by the BERT models. The LLM biases, however, were often opposite to the BERT models, which typically encoded the biases inherent in the fine-tuning population. Rule-based methods were consistently the least biased, and some of the LLMs were very biased, in particular those based on Qwen v 2.5 and 3.

### 1.4.5 Economic Comparison

A complete economic analysis was undertaken by model, as per ***Table***.

**Table 2:**
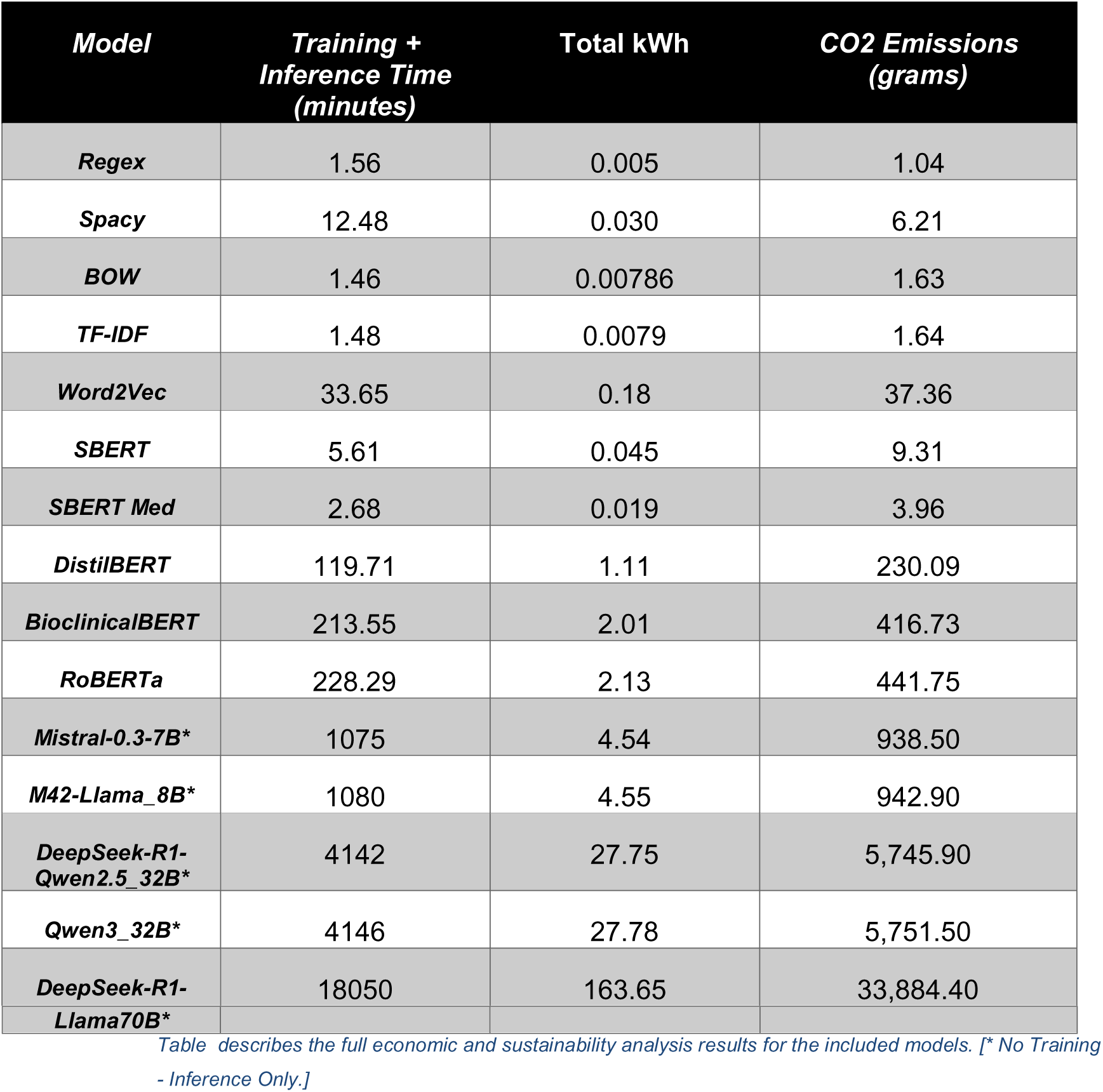
Full Economic Results By Model.

Fine-tuning BERT models is moderately costly in terms of computation time, but inference is comparatively rapid. SBERT models are lightweight and can be faster than even running Spacy phrasematcher pipelines, yielding better results. Regex, BOW and TF-IDF take the crown for speed and efficiency at this task. Running inference with LLMs is slow, and, in the case of the largest models, it is prohibitively costly, particularly when the degree of associated performance gain is considered.

### 1.4.6 Explainability Analysis

The string-based methods are entirely explainable. SHAP plots for BOW, TF-IDF, and Word2Vec all highlight similar word token patterns, as highlighted in *Error! Reference source not found.*. SHAP plots for distilBERT suggest that the tokens ‘micoscopic’, ‘mapping’, and ‘biopsy’ were positively weighted in the model, while bioclinicalBERT emphasised ‘mapping’, ‘reporting’, and ‘terminal’. RoBERTa, however, weighted ‘specialist’, ‘colon’, ‘pain’, ‘inflammatory’, and ‘external’, indicating that this model developed a different understanding of the data than the other two BERT models. These features are prevalent in the dataset, likely contributing to the model’s collapse, as it predicts every instance as IBD. LIME analysis of the same models assigned negative weights to terms like ‘bowel’ and ‘biopsies’. Examining one of these examples visually, as per *Error! Reference source not found.,* proves to be illuminating as it demonstrates how sensitive transformers are to self-contradictory information in a clinical record.

LLM performance was overall much the same, with contradictory information and statements, such as ‘possible’ IBD, which often confused them. In several instances, the LLMs did not realise that microscopic colitides were forms of IBD. This aspect of LLM explainability will be delved into in much more detail in a future study.

## 1.5 Discussion

This study has highlighted numerous challenges in identifying clinical IBD patient cohort groups through NLP, many of which have not been previously documented(5,6,48). The study emphasises that patient-level cohort identification is somewhat context-dependent and cannot be conducted in isolation. BOW and some types of BERT models (SBERT, distilBERT, and bioclinicalBERT) proved to be the most effective; however, other algorithms, like LLMs, demonstrate useful emergent qualities even if fairness and cost limit their scale. This example highlights, in a microcosm, one of the considerable problems with foundation models and why they cannot be used safely in a healthcare context without further development work. In particular, Qwen 2.5 and 3 are shown to be substantially biased. Simple rule-based methods are by far the most effective at maximising recall and have the added advantages of low bias and cost while remaining fully interpretable.

It should be simple for others to implement the models in other English-speaking contexts now that they are released open source, because very limited pre-processing is required. A simple set of <u>demo apps</u> is made available for users to demonstrate how easy the models are to use. The models should function correctly whether PII is redacted or not, and they should operate with either the transformers or scikit-learn libraries without difficulty. The LLM framework is proven to generalise in this study; however, the generalisability of other models remains to be established.

Fairness is an issue for all types of models, except for rule-based classifiers. Locally trained models reflect the biases of the local population, while LLMs typically contain opposing but generally more extreme biases. Model collapse was an issue with some models, such as RoBERTa, suggesting that pre-training size alone is not particularly beneficial. Weaknesses of this study include class imbalance within the high-yield, high-information-availability training and validation cohort. Many of these issues are mitigated through the use of the F1 score and MCC as primary evaluation metrics. The sample size, methodology, and low levels of missingness are strong features of this study. Another potential criticism of the study is the use of 8-70B parameter LLMs due to hardware restrictions. While there is a slight chance that the ‘full’ R1, Mistral, or Qwen model would have performed significantly better, the evidence we have suggests that large LLMs are, in fact, even less faithful when managing factual information, even if readability and informativeness improve(49). A subsequent study is planned to inspect the quality of LLMs at this task.

The strengths of this study include the level of detail provided in the analysis, the transparent reporting methods employed, and the open sharing of source code and models, which is essential for substantial progress being made in this field(5). The validation of the gold standard cohort in this study was extremely robust and led by a senior gastroenterologist with strong informatics experience. Unfortunately, since the data was not consented to, it cannot be shared at present. One of the main weaknesses of this study was the inclusion of patients with microscopic colitis in the IBD cohort. Technically speaking, microscopic colitis is not IBD, even if it might be a precursor(50). This was added for local service reasons. However, if there is strong demand for it, then the models will be retrained without this inclusion – a relatively simple undertaking. This highlights another major weakness of this study, namely its single-site nature. Conducting multi-site studies like this isn’t easy due to the localised nature of high-performance computing (HPC) infrastructure. Hopefully, with the arrival of the secure data environments (SDEs) this will soon change(51).

Other authors do not seem to be aware of the document-to-patient mapping problem(52–57), with most studies primarily only focusing on extracting clinical concepts from annotated free-text clinical records. Future planned work will explore optimal methods to enhance LLM performance in cohort identification and document classification tasks and assess hybrid model-based methods to identify clinical cohorts successfully.

## 1.6 Conclusion

Older NLP technologies from over 50 years ago, such as BOW, can outperform even the most modern NLP technologies, such as Deepseek R1, when properly trained to detect clinical cohorts. Although they don’t require pre-training, LLMs are cumbersome, slow and expensive to use for cohort identification tasks and tend towards low recall.

All the models trained in this study are made open source/weight to benefit patients and the broader research community.

### 1.6.1 Published Declarations Alongside Manuscript

**Twitter:** Matt Stammers: @MattStammers_

## Author Contributors

MS performed all analyses and prepared the final data. MG, RN, CM, and JB (MS’s supervisors) provided critical feedback regarding the manuscript. **MS** is the primary guarantor for the review and the corresponding author.

## Acknowledgements

The local SETT data and AI, CIRU, and Gastroenterology/IBD teams are acknowledged for building the wider infrastructure that made this project possible.

## Patient consent for publication

Not applicable

## Data Availability

Data sharing is not possible in this study because the data were not collected for this purpose; however, secondary data can be made available upon request.

## Code Availability

All codes used in the analytics for this project are made available open source on GitHub at https://github.com/MattStammers/An_Open_Source_Collection_Of_IBD_Cohort_Identification_Models

## Model Availability

Models are made fully accessible open source at: https://huggingface.co/collections/MattStammers/a-collection-of-ibd-bert-models-682b01badbaa646380f54b14 and in the <u>GitHub</u> repo. The LLM’s are all open source and can be accessed at the following links:

1. **Mistral 7b-v0.3-Instruct**(58): https://huggingface.co/mistralai/Mistral-7B-Instruct-v0.3
2. **Llama3-Med42-8b**(59): https://huggingface.co/m42-health/Llama3-Med42-8B
3. **Deepseek-Qwen2.5-32B**(60): https://huggingface.co/deepseek-ai/DeepSeek-R1-Distill-Qwen-32B
4. **Qwen-v3-32**(61): https://huggingface.co/Qwen/Qwen3-32B
5. **Deepseek-Llama-70B**(62): https://huggingface.co/deepseek-ai/DeepSeek-R1-Distill-Llama-70B

## Patient and Public Involvement

An IBD patient from our local patient panel contributed to the development of the ethics application and study protocol.

## Human Ethics and Consent to Participate Statement

The Wessex REC and HRA provided research ethics board approval for this study (IC-IBD-23/SC/0152) on 16/05/2023 (https://www.hra.nhs.uk/planning-and-improving-research/application-summaries/research-summaries/ic-ibd-ibd-cohort-identification-study/).

