## Appendix B for "From Rule-Based to DeepSeek R1 – A Robust Comparative Evaluation of Fifty Years of Natural Language Processing (NLP) Models To Identify Inflammatory Bowel Disease Cohorts"

Appendix B: Platform Setup and Usage of UMLS for Text Normalisation

##### Environment & API Setup

This study was conducted within the hospital using four locally deployed L40S graphics cards housed in a bare-metal server. The LLM server was set up using Gradio 1.5.1(60), accessed through a local HTTP-based API connection with exponential backoff to avoid overwhelming the system, an asynchronous queue, and a .json filter to check for correctly formatted responses. The platform was developed using Python 3.11 and the torch, transformers, and accelerate libraries.

##### Dev Ops, Code & Model Sharing

All constants and filters were governed by a standard configuration, with local caching where appropriate. Tests guaranteed platform stability, and a common set of utilities reduced the risk of error.

The code and the trained models are provided in full. The code is available on GitHub: <https://github.com/MattStammers/An_Open_Source_Collection_Of_IBD_Cohort_Identification_Models> under a CC-BY-NC 4.0 license, and the transformer models are made openly available on Huggingface (an open-source language model site) as part of a collection: <https://huggingface.co/collections/MattStammers/a-collection-of-ibd-bert-models-682b01badbaa646380f54b14> under the same license.

##### UMLS Standardisation vs Free-Text

The Unified Medical Language System (UMLS)(61) metathesaurus provides a mapping structure among various medical vocabularies and serves as a comprehensive thesaurus and ontology of biomedical concepts. Each fully trained model was run with and without UMLS text standardisation utilising a filtered version of MRCONZO.rrf to identify only IBD-specific drug concepts to investigate whether this tool enhances or impedes clinical performance during this clinical cohort identification task. These synonymous concepts were linked to their corresponding preferred term (PT), which was then substituted in the text. The associated concept unique identifiers (CUI’s) (some examples of these are given in ***Appendix B)*** were omitted from the text to prevent the model from learning inappropriately from them. Only English non-obsolete terms from all source vocabularies (ontologies) were employed in the form of the PT. A decision was made at the outset that the best-performing form of the data would be used for the evaluation.

***UMLS Normalised IBD Terms***

**Normalised IBD Predictor Terms**

Normalised IBD-predictor terms identified in the training set included: ["Ulcerative Colitis", "Crohn's Disease", "IBD", "Inflammatory Bowel Disease”, "Proctitis", "Collagenous Colitis", "Microscopic Colitis", "Lymphocytic Colitis"].

**Normalised IBD Drug Predictor Terms**

Normalised IBD-drug predictor terms included: ["Prednisolone", "Octasa", "Salofalk", "Pentasa", "Asacol", "Mesalazine", "Budenofalk", "Budesonide", "Sulfasalazine", "Salazopyrin", "Tofacitinib", "Adalimumab", "Methotrexate", "Etanercept", "Azathioprine", "Mercaptopurine", "Ustekinumab", "Infliximab", "Vedolizumab", "Mirikizumab", "Upadacitinib", "Filgotinib”].

**Cuis for Normalised Drug Terms**

- Term: mercaptopurine, CUI: ['C0000618']
- Term: azathioprine, CUI: ['C0004482']
- Term: hydrocortisone, CUI: ['C0020268']
- Term: methotrexate, CUI: ['C0025677']
- Term: prednisolone, CUI: ['C0032950']
- Term: sulfasalazine, CUI: ['C0036078']
- Term: budesonide, CUI: ['C0054201']
- Term: mesalazine, CUI: ['C0127615']
- Term: infliximab, CUI: ['C0666743']
- Term: salofalk, CUI: ['C0678169']
- Term: pentasa, CUI: ['C0678171']
- Term: asacol, CUI: ['C0678172']
- Term: etanercept, CUI: ['C0717758']
- Term: adalimumab, CUI: ['C1122087']
- Term: ustekinumab, CUI: ['C1608841']
- Term: vedolizumab, CUI: ['C2742797']
- Term: tofacitinib, CUI: ['C2930696']
- Term: salazopyrin, CUI: ['C4082693']
- Term: filgotinib, CUI: ['C4310256']
- Term: upadacitinib, CUI: ['C4726929']
- Term: mirikizumab, CUI: ['C5139904'**]**
