## Appendix C for "From Rule-Based to DeepSeek R1 – A Robust Comparative Evaluation of Fifty Years of Natural Language Processing (NLP) Models To Identify Inflammatory Bowel Disease Cohorts"

Appendix C: LLM Templating System

These LLM responses underwent an identical analytics process to the other models, using the same threshold of >= five as the cutoff for diagnosis. This method helped mitigate some of the ‘black-box’ problems associated with LLMs by mandating a human-interpretable, quantifiable, structured response. It also allowed a set of discrete probabilities to be returned, facilitating a slightly crude but nonetheless better-than-nothing assessment of model calibration.

Below is the core prompt template (See ***Figure C***) derived through extensive experimentation and testing, this prompt is based on the experiences developed during Microsoft^TM^’s learnings during the construction of MedPrompt(19), and is only 409 tokens long, leaving ample space for dynamic content (the context window limit is set at 4,000 for these experiments to provide an entirely level playing field. Building on MedPrompt, this prompt also leverages the Clue and Reasoning Prompt (CARP) method(31), which is particularly effective in document text classification tasks using LLMs. In-context learning (ICL) is intentionally not used here to allow for zero-shot performance assessment for this clinical task.

The prompt establishes the context, requests clues, reasoning, and the outcome in JSON format, provides a chain of thought (CoT), and then presents the specific contextual information as input, followed by a reiteration of the instructions. The prompt is sent to non-thinking models at a temperature of 0.75 (M42/Mistral) and 0.6 for the thinking models (Deepseek/Qwen). These settings were selected based on the manufacturer's instructions.

***Figure C*:** *Zero-Shot Prompt Base Template:*

combined_message = f"""

As a resident / junior doctor tasked with analysing gastroenterology reports. Your task is to extract key information and provide a summary in JSON format.

**Instructions:**

1. **CLUES:**

Describe any important insights or motivations based on the reports. Keep it brief and relevant.

2. **REASONING:**

Explain the reasoning behind the diagnostic conclusion.

3. **OUTCOME:**

Provide the outcome in JSON format with the following fields:

- **Title:** A summary title for the diagnosis.

- **Features:** A list of key features extracted from the reports.

- **Likelihood of IBD:** A rating from 0 to 10 (lowest to highest) indicating the likelihood of Inflammatory Bowel Disease (IBD).

- **Certainty Level:** A rating from 0 to 10 (lowest to highest) indicating the certainty of the diagnosis.

- **Complexity of Case:** A rating from 0 to 10 (lowest to highest) indicating the complexity of the case.

**Please provide your response in the following format:**

**CLUES:**

<CLUES>

**REASONING:**

<REASONING>

**OUTCOME:**

```json

{{

"Title": "<Title>",

"Features": [

"<Feature 1>",

"<Feature 2>"

],

"Likelihood of IBD": <Likelihood of IBD>,

"Certainty Level": <Certainty Level>,

"Complexity of Case": <Complexity of Case>

}}

```

**INPUT:**

**PRECEDING CLINIC LETTER:**:

{preceding clinic_letter} # Dynamic Element

**ENDOSCOPY REPORT:**:

{endo_report}	# Dynamic Element

**HISTOPATHOLOGY REPORT:**:

{hist_report}	# Dynamic Element

**FOLLOWING CLINIC LETTER:**:

{following clinic_letter} # Dynamic Element

**Important:** Provide only a SINGLE JSON response regarding the likelihood of IBD. Do not repeat or include multiple JSON blocks. Please ensure the final response is in the correct JSON format.

"""

*Figure C above contains the zero-shot prompt for LLM prompting used in the study. It describes the Chain of Thought (CoT) utilised in the study, as well as prompting the LLM to provide reasons for its responses, finally returning the answer in .json format.*

The task is to rate each document out of 10 based on the dimensions of likelihood, certainty, and complexity. This compels the LLM to articulate its thought process and assign a numerical weight to its decisions. Subsequently, this weight can be utilised as a proxy measure of LLM quality, confidence, and optimism, all while providing a quantifiable measure of IBD likelihood.

For these tests, the dynamic content is given in chronological order in each application programming interface (API) call with no knowledge/memory of the prior call. If a malformed .json object was received, a simple attempt was made to repair common errors to maximise the response rate without changing the response. However, if this fails, the response is discarded as invalid.
