## Appendix D for "From Rule-Based to DeepSeek R1 – A Robust Comparative Evaluation of Fifty Years of Natural Language Processing (NLP) Models To Identify Inflammatory Bowel Disease Cohorts"

Appendix D: Metrics of Interest

***Table D:*** *Performance Metrics Used in this Study*

| *Term* | *Description* |
| --- | --- |
| Accuracy | The percentage of results that were correct among all results from the system. Calc: (TP+TN)/(TP+FP+TN+FN). |
| Precision (PPV) | Also called positive predictive value (PPV), it is the percentage of true positive results among all results that the system flagged as positive. Calc: TP/(TP+FP). |
| Negative Predictive Value (NPV) | The percentage of results that were true negative (TN) among all results that the system flagged as negative. Calc: TN/(TN+FN). |
| Recall | Also called sensitivity. The percentage of results flagged positive among all results should have been obtained. Calc: TP/(TP+FN). |
| Specificity | The percentage of results that were flagged negative among all negative results. Calc: TN/(TN+FP). |
| F1-Score | In this case, the harmonic mean of PPV/precision and sensitivity/recall is unweighted. Calc: 2 × (Precision x Recall)/(Precision + Recall). Moderately useful in the context of class imbalance. |
| Matthews’ Correlation Coefficient (MCC) | A statistical measure is used to evaluate the quality of binary classifications. Unlike other metrics, MCC considers all four categories of a confusion matrix. Calc: MCC=(TP×TN)−(FP×FN)√(TP+FP)(TP+FN)(TN+FP)(TN+FN) |
| Precision / Recall AUC | Represents the area under the Precision-Recall curve, which plots Precision against Recall at various threshold settings. It is more resistant to class imbalance than alternatives like AUROC, which is why it is used here. |
| Demographic Parity (DP) | Demographic Parity, also known as *Statistical Parity*, requires that the probability of a positive prediction is the same across different demographic groups. Calc: DP = P(Y^=1∣A=a)=P(Y^=1∣A=b)(35). where Y^ is the predicted outcome, and A represents the protected attribute (e.g., race, gender). This figure is given as an absolute difference where +ve values suggest the more privileged group gains and negative the reverse. |
| Equal Opportunity (EO) | Equal Opportunity focuses on equalising the true positive rates across groups. That is, among those who truly belong to the positive class, the model should predict positive outcomes at equal rates across different groups. Calc: EO = P(Y^=1∣Y=1,A=alpha)=P(Y^=1∣Y=1,A=b)(36). Where Y^ is the predicted outcome, and Y is the actual outcome. If there is no bias, then the value will be equal for all groups. A higher value indicates a bias against the group considered more vulnerable. |
| Disparate Impact (DI) | Divides the protected group’s positive prediction rate by that of the most-favoured group; if the ratio is below 0.8 (or above 1.25), disparate impact is considered present. Calc: DI=P(Y^=1∣A=unfavoured)P(Y^=1∣A=favoured)(37). If there is no significant bias, this value will be 0.8-1.25. A higher value indicates a bias against the group considered more vulnerable. |
| Execution Time/Energy/CO2 Emissions | Measured in minutes and total energy consumption in kilowatt-hours (kWh), which can then be extrapolated to CO2 emissions via a conversion factor set at 0.20705 Kg CO_2_e per kWh for this study(39). |

*A* full set of measurement metrics are used in this study as highlighted in ***Appendix D***.

*D above provides a complete list of the metrics used in this study. The non-expounded abbreviations include: TP = True Positive, FP = False Positive, FN = False Negative, TN = True Negative.*
