## Appendix E for "From Rule-Based to DeepSeek R1 – A Robust Comparative Evaluation of Fifty Years of Natural Language Processing (NLP) Models To Identify Inflammatory Bowel Disease Cohorts"

Appendix A Model Handling

#### Fully Trained Models

##### Rule-Based (RB) Models

This study employed two rule-based approaches: a simple regex and a spacy model featuring negation.

###### Simple Regex

A simple regex-based string-search model was developed to find the terms as described in the code. The Python built-in regex module ‘re’ detected IBD and drug terms, ignoring cases, and predicted a ‘1’ if a match was found and a ‘0’ if not.

The modular components simulate training for the rule-based pipelines to ensure that the standard approach operates correctly across all the modules. Each pipeline integrates all the fairness, economic, and explainability components modularly. However, explainability analyses do not particularly apply to this and the spacy pipeline, as they are inherently explainable.

###### Spacy with Negation

A spacy NLP pipeline enhanced with the negspacy ‘negex’ component was created to handle text negations. Negation terms were derived following a manual document review and are provided in ***Appendix F***. ‘en_core_web_md’ was used as the core spacy language model because scientific models such as those by scispacy(62) did not appear to improve performance and incurred significant module backward compatibility problems so were discarded.

A phrase matcher pipeline was implemented to enhance the efficiency of bulk matching. Normalised scores were generated based on the number of matches for IBD, with the final threshold set at >=1 for highly sensitive detection, as increasing the threshold did not significantly improve precision but did reduce recall. Negation was maintained during the pipeline's usage.

##### Machine Learning (ML) Models

These models create different word vectors, which were fed into an LR classifier. In each instance, classification thresholds were set at 0.5.

###### Bag of Words (BOW) Model

This BOW word vector model weighs documents using a CountVectorizer and then fits an LR binary classifier to predict ‘IBD’ or ‘Not-IBD’. These models disregard word order and context, focusing primarily on word multiplicity(7). Model training and inference are swift. This facilitated numerous experiments with this model type. Initially, default ridge (L2) regression was used along with a limited-memory Broyden-Fletcher-Goldfarb-Shanno (lbfgs)(63) solver algorithm, but despite being quick, this yielded only mediocre results. Lasso (L1) regression with a liblinear solver(64), however, worked far better. This is probably because the high-dimensional space in the clinical text is overly complex meaning that feature reduction improved performance. Tree models, such as XGBoost, did not perform better than LR for this task and were substantially slower. Eventually, gridsearchCV(65) was added with 1,000 iterations, a mixed penalty, and an AUROC scorer seeded 42. This performed exceptionally well therefore retained as the base setup for the majority of the ML algorithms. These models were all saved in .joblib format to make them easier to deploy than they would be as pickle (.pkl) files.

###### Term Frequency-Inverse Document Frequency (TF-IDF) Model

This vector model operates by identifying words that are significant in a document but not widespread across the collection of documents(9). Consequently, it is less susceptible to common-word distortion and frequently represents a more robust text-mining algorithm than BOW, particularly in more complex classification tasks. The best results were obtained with a setup similar to the BOW pipeline, so this is how it was configured. All the code is provided for complete transparency and to enable others to replicate or improve upon the experiment.

###### Spacy(Word2Vec)

This model employs Spacy’s en_core_web_md to generate word vectors using a custom Word2VecTransformer(), which also feeds into an LR classifier. Gensim was not used because it was incompatible with the analytics environment. As these models are only embedded in a 300-dimensional vector space further tuning the hyperparameters did not significantly improve performance unlike with SBERT.

#### Finetuned Transformers

##### SBERT Vector Model

A custom sci-kit learn transformer was created for these Sentence-BERT (SBERT) pipelines – SbertEmbeddingTransformer() using the ‘all-mpnet-base-v2’ base model(66), because this was built upon a considerable corpus of datasets(67), known to demonstrate superior embedding performance, mapping to a 768-dimensional dense vector space. However, it should be noted that the datasets this model was trained on are primarily from Reddit, stack exchange, and wikis—not from clinical sources. The SBERT models were stored both in joblib and onnx transformer format to make them cross-compatible.

##### SBERT Med Vector Model

As ‘all-mpnet-base-v2’ is not pre-trained for clinical applications, a biobert-based SBERT vector embedding model was also tested(45). This model was trained with an Adam optimiser over the Stanford Natural Language Inference (SNLI)(68), Multi-Genre Natural Language Inference (MNLI)(69), Scientific Natural Language Inference (SCINLI)(70), Scientific Tail (SCITAIL)(71), Medical Natural Language Inference (MEDNLI)(72) and Semantic Textual Similarity Benchmark (STSB)(73) datasets.

However, unlike the base SBERT implementation, this model did not benefit from further hyperparameter tuning, so gridsearchCV was turned off for this model’s pipeline.

##### DistilBERT

DistilBERT was created by Hugging Face in 2019(15) as a BERT option for resource-constrained environments. It is 40% smaller than the original BERT model, which had 110 million parameters(13), yet it retains 97% of the performance characteristics and operates 60% faster. DistilBERT has its own tokeniser and sequence classification handlers. Captum was used to explore integrated gradients along with LIME(42) text explorers conducted over a small selection (n=5) of each document type. Many of the SHAP and LIME processes were transferred to the CPU to prevent memory overspills, as L40S units only have 48GB of VRAM.

Training for the distilBERT models was conducted over 20 epochs, with convergence usually occurring after approximately 15 epochs, when the learning rate was set at 0.0005. Training progressed through the same pattern as for the other models – document model training and then validation, followed by document-to-patient validation and then finally, patient-level model training and validation. Again, classifiers were seeded 42. These models were all saved as full safe tensor transformers and uploaded to [Hugging Face](https://huggingface.co/collections/MattStammers/a-collection-of-ibd-bert-models-682b01badbaa646380f54b14) after training.

##### BioClinicalBERT (BioClinicalBERT)

BioClinicalBERT(17) is a larger fine-tuned BERT model initialised from BioBERT(74) and trained on 880 million note-event words from MIMIC III(75). Unlike Distilbert, this model is several gigabytes (GB) in size and requires significantly longer to fine-tune. It employs a standard BERT Tokeniser; otherwise, the pipeline and model handling were nearly identical to DistilBERT’s, with the same learning rate, batch size and number of epochs.

##### RoBERTa

RoBERTa stands for Robustly Optimised BERT. The original version of BERT(16) was trained on several additional web corpora. However, the creators of RoBERTa felt that the original BERT model was under-trained, so they added additional pre-training. ‘Biomed_roberta_base’(76) was the RoBERTa model selected for this study because it was subsequently trained on 2.68 million scientific papers from Semantic Scholar, adding 7.55 billion (B) tokens to the model(46), making the largest BERT model tested in this study with some degree of scientific pre-training but not specifically clinical pre-training. RoBERTa has a specific RobertaTokeniser() and SequenceClassification handler.

#### Prompt Engineered Transformers

These GPT models selected as the best open-source pure language models available from July 2024 to June 2025 to test the study's assumptions, allowing for over a year’s worth of evolution in the field and a variety of model sizes to be tested.

They range from relatively small (Mistral-Instruct-0.3-7B), which requires approximately 15GB VRAM to deploy locally and can be loaded onto a decent laptop, to relatively large ‘thinking’ models like Deepseek-R1-Distill-Llama3-70B, which necessitates at least 140GB VRAM for local deployment.

All the models were deployed on the same 4*L40S GPUs (mid-level commercial-grade NVIDIA GPU chipset) to maintain a level playing field and allow for direct performance comparison without necessitating quantisation at any point.

##### Mistral-Instruct-0.3-7B

This instruct-tuned model(55), released in July 2024 by Mistral^TM^, a French-American startup, was chosen as a robust, light, general-purpose LLM due to its extensive vocabulary. At the time testing commenced it was the most advanced small open-source model available. It features a 4.8k token context window (which at that time was unusual for models of this size) and can be deployed with under 15GB of unquantised VRAM.

In all of these pipelines, a score of 5 or greater for IBD was deemed suggestive of IBD. This cutoff was selected to reflect the standard threshold of 0.5, to maintain a level playing field across all the models.

##### M42-Health/Llama3-8B

This model was released in August 2024 by m42 HealthTM, a tech-enabled healthcare company based in the UAE. It was trained on top of Llama3-8B(56). It was chosen because it was one of the first open-source medically trained LLMs that functioned correctly as a small instruct-tuned model. As recommended, both the smaller models were run at a temperature of 0.75.

##### Deepseek-R1-Distill-Qwen-2.5-32B

Deepseek R1(20) was released in January 2025 by the Chinese startup Deepseek^TM^ and created a stir when people realised it was nearly as capable as ChatGPT^TM^ o1, yet open-source. Papers are already beginning to emerge suggesting that these models are competitive in performance for clinical applications(77).

Deepseek R1 was initially trained through large-scale reinforcement learning (RL) using a type of proximal policy optimisation(78) called group relative policy optimisation (GRPO) to assist in solving mathematics puzzles(79). To enhance the model's readability, cold-start data and a multi-stage training pipeline with additional reinforcement learning (RL) were incorporated to reach the R1 checkpoint without requiring human supervision during training. This granted the model the ability to ‘think’ before answering. To thoroughly assess these new ‘thinking’ open source LLMs, the same experiment was conducted with Deepseek R1 Qwen 32B, Qwenv3 32B, and Deepseek R1 Llama 70B, all set at a temperature of 0.6.

##### Qwen3-32B

Qwen3-32B(23) was released by BaiduTM in April 2025 and is the most up-to-date of the models tested. It is fluent in over 100 languages. The complete model comprises 235B parameters, with this 32B version being the second largest. Comparing it to Deepseek’s versions indirectly compares Qwen v2.5 and 3.

Llama4(80) is notably multimodal and, therefore, unsuitable for this experimental setup, which centres on the cutting-edge capabilities of pure language models, which don’t tend to be as associated with lessened computational efficiency(81) and hidden performance defects(82).

##### Deepseek-R1-Distill-Llama3-70B

This is the largest model tested. Deepseek^TM^ released it in January 2025. Its performance metrics are almost equivalent to the full R1 671B model(20). This model was selected because it is built on Llama3 70B(83), which is Meta^TM^'s most advanced US pure language foundation model. U these distilled versions facilitates, to a lesser degree, direct comparisons between the performance of the underlying foundation models, namely Qwen, Mistral, and Llama, at this task.
