## Appendix F for "From Rule-Based to DeepSeek R1 – A Robust Comparative Evaluation of Fifty Years of Natural Language Processing (NLP) Models To Identify Inflammatory Bowel Disease Cohorts"

Appendix F: Negation Termsets

Below is the negation term set dictionary used for model 2: spacy with negation

```
NEG_TERMSET = {
        "pseudo_negations": [
            "without",
            "no",
            "negative for",
            "rule out",
            "exclude",
            "absence of"
        ],
        "preceding_negations": [
            "not",
            "no evidence of",
            "without",
        ],
        "following_negations": [
            "no evidence of",
            "no signs of",
            "no history of",
            "no indication of"
        ],
        "termination": [
            ";",
            "."
        ]
    }
```
