## Appendix G for "From Rule-Based to DeepSeek R1 – A Robust Comparative Evaluation of Fifty Years of Natural Language Processing (NLP) Models To Identify Inflammatory Bowel Disease Cohorts"

Appendix G: Demographic Results

***Table G:*** *Training and Validation Cohort Metrics*

| *Cohort* | *Feature* |  |  |  |  |  | *p-value* |
| --- | --- | --- | --- | --- | --- | --- | --- |
| *Continuous Variables* |  | ***Mean*** | ***Median*** | ***IQR*** | ***Skewness*** | ***Kurtosis*** |  |
| *Training* | *Age* | *44.21 +/- 95%CI: 0.81* | *42.27* | *29.15* | *0.2996* | *-0.9797* | *0.2973* |
| *Validation* | *Age* | *45.01 +/- 95%CI: 1.27* | *42.91* | *30.78* | *0.2497* | *-1.1526* | *0.2973* |
| *Training* | *IMD* | *4.89 +/- 95%CI: 0.13* | *4.00* | *4.00* | *0.2701* | *-1.0418* | *0.9942* |
| *Validation* | *IMD* | *4.89 +/- 95%CI: 0.19* | *4.00* | *4.00* | *0.3348* | *-0.8957* | *0.9942* |
| *Discrete Variables* |  | ***Proportion*** |  |  |  |  |  |
| *Training* | *Gender (Female)* | *50.65%* |  |  |  |  | *0.0148* |
| *Validation* | *Gender (Female)* | *53.52%* |  |  |  |  | *0.0148* |
| *Training* | *Ethnicity (White)* | *81.91%* |  |  |  |  | *0.0002* |
| *Validation* | *Ethnicity (White)* | *79.56%* |  |  |  |  | *0.0002* |

*Table describes the cohort statistics for the training and validation cohorts*

No information on age, IMD, or gender data was missing. However, n=174 (10.8%) of patients were not asked about their ethnicity.
