## Appendix H for "From Rule-Based to DeepSeek R1 – A Robust Comparative Evaluation of Fifty Years of Natural Language Processing (NLP) Models To Identify Inflammatory Bowel Disease Cohorts"

Appendix H: Document Level Results

***Table H:*** *Full Model Results (Document-Level)*

| *Model* | *Doc Coverage* | *Accuracy* | *Precision* | *Recall* | *Specificity* | *NPV* | *F1 Score* | *MCC* |
| --- | --- | --- | --- | --- | --- | --- | --- | --- |
| *Fully Trained* | | | | | | | | |
| *Regex* | *768 (100.00%)* | *84.76% (CI: 81.98% - 87.18%)* | *85.02% (CI: 82.08% - 87.55%)* | *97.54% (CI: 95.92% - 98.53%)* | *40.61% (CI: 33.41% - 48.23%)* | *82.72% (CI: 73.05% - 89.42%)* | *90.85% (CI: 89.24% - 92.46%)* | *0.5083 (CI: 0.4232 - 0.5939)* |
| *Spacy* | *768 (100.00%)* | *83.46% (CI: 80.67% - 85.92%)* | *83.54% (CI: 80.67% - 86.05%)* | *98.86% (CI: 97.67% - 99.45%)* | *21.05% (CI: 15.33% - 28.21%)* | *82.05% (CI: 67.33% - 91.02%)* | *90.56% (CI: 89.81% - 91.40%)* | *0.3614 (CI: 0.2764 - 0.4453)* |
| *BOW* | *768 (100.00%)* | *89.58% (CI: 87.22% - 91.55%)* | *89.30% (CI: 86.75% - 91.40%)* | *98.86% (CI: 97.67% - 99.45%)* | *51.97% (CI: 44.08% - 59.77%)* | *91.86% (CI: 84.14% - 96.00%)* | *93.84% (CI: 92.82% - 94.89%)* | *0.6423 (CI: 0.5742 - 0.7119)* |
| *TF-IDF* | *768 (100.00%)* | *83.85% (CI: 81.08% - 86.29%)* | *83.42% (CI: 80.57% - 85.94%)* | *99.68% (CI: 98.82% - 99.91%)* | *19.74% (CI: 14.19% - 26.78%)* | *93.75% (CI: 79.85% - 98.27%)* | *90.83% (CI: 90.21% - 91.53%)* | *0.3871 (CI: 0.3133 - 0.4572)* |
| *Word2Vec* | *768 (100.00%)* | *82.16% (CI: 79.30% - 84.71%)* | *81.81% (CI: 78.89% - 84.40%)* | *100.00% (CI: 99.38% - 100.00%)* | *9.87% (CI: 6.07% - 15.64%)* | *100.00% (CI: 79.61% - 100.00%)* | *89.99% (CI: 89.53% - 90.52%)* | *0.2841 (CI: 0.2065 - 0.3537)* |
| *Finetuned* | | | | | | | | |
| *SBERT* | *768 (100.00%)* | *90.36% (CI: 88.07% - 92.26%)* | *90.45% (CI: 87.99% - 92.45%)* | *98.38% (CI: 97.04% - 99.12%)* | *57.89% (CI: 49.95% - 65.45%)* | *89.80% (CI: 82.23% - 94.36%)* | *94.25% (CI: 93.19% - 95.17%)* | *0.6720 (CI: 0.6075 - 0.7345)* |
| *SBERT Med* | *768 (100.00%)* | *83.72% (CI: 80.95% - 86.17%)* | *83.68% (CI: 80.82% - 86.18%)* | *99.03% (CI: 97.89% - 99.55%)* | *21.71% (CI: 15.90% - 28.92%)* | *84.62% (CI: 70.27% - 92.75%)* | *90.71% (CI: 89.96% - 91.48%)* | *0.3763 (CI: 0.2959 - 0.4530)* |
| *DistilBERT* | *768 (100.00%)* | *91.93% (CI: 89.79% - 93.65%)* | *92.10% (CI: 89.78% - 93.92%)* | *98.38% (CI: 97.04% - 99.12%)* | *65.79% (CI: 57.94% - 72.86%)* | *90.91% (CI: 84.07% - 94.99%)* | *95.13% (CI: 94.14% - 96.20%)* | *0.7298 (CI: 0.6624 - 0.7898)* |
| *BioclinicalBERT* | *768 (100.00%)* | *90.76% (CI: 88.50% - 92.61%)* | *90.73% (CI: 88.30% - 92.70%)* | *98.54% (CI: 97.25% - 99.23%)* | *59.21% (CI: 51.26% - 66.70%)* | *90.91% (CI: 83.62% - 95.14%)* | *94.47% (CI: 93.54% - 95.44%)* | *0.6866 (CI: 0.6225 - 0.7490)* |
| *RoBERTa* | *768 (100.00%)* | *91.28% (CI: 89.07% - 93.07%)* | *91.78% (CI: 89.43% - 93.65%)* | *97.89% (CI: 96.42% - 98.76%)* | *64.47% (CI: 56.59% - 71.64%)* | *88.29% (CI: 80.99% - 93.03%)* | *94.74% (CI: 93.72% - 95.74%)* | *0.7066 (CI: 0.6434 - 0.7671)* |
| *Prompt Engineered* | | | | | | | | |
| *Mistral-0.3-7B* | *768 (100.00%)* | *81.66% (CI: 80.48% - 82.78%)* | *89.47% (CI: 88.26% - 90.56%)* | *82.85% (CI: 81.45% - 84.16%)* | *79.12% (CI: 76.91% - 81.18%)* | *68.30% (CI: 65.99% - 70.53%)* | *86.03% (CI: 85.09% - 86.97%)* | *0.5983* |
| *M42-Llama_8B* | *768 (100.00%)* | *80.77% (CI: 79.58% - 81.91%)* | *88.51% (CI: 87.28% - 89.65%)* | *82.45% (CI: 81.04% - 83.77%)* | *77.21% (CI: 74.94% - 79.33%)* | *67.37% (CI: 65.04% - 69.62%)* | *85.37% (CI: 84.39% - 86.39%)* | *0.5774* |
| *DeepSeek-R1- Qwen2.5_14B* | *768 (100.00%)* | *83.92% (CI: 82.78% - 85.00%)* | *88.70% (CI: 87.48% - 89.81%)* | *87.65% (CI: 86.39% - 88.80%)* | *75.88% (CI: 73.51% - 78.10%)* | *73.99% (CI: 71.60% - 76.25%)* | *88.17% (CI: 87.25% - 89.07%)* | *0.6311* |
| *DeepSeek-R1- Qwen2.5_32B* | *768 (100.00%)* | *84.21% (CI: 80.20% - 87.53%)* | *91.03% (CI: 86.67% - 94.06%)* | *84.52% (CI: 79.54% - 88.47%)* | *83.59% (CI: 76.22% - 89.01%)* | *73.29% (CI: 65.58% - 79.80%)* | *87.65% (CI: 84.39% - 90.46%)* | *0.6619* |
| *Qwen3_32BM* | *768 (100.00%)* | *81.51% (CI: 77.32% - 85.07%)* | *92.56% (CI: 88.25% - 95.37%)* | *78.35% (CI: 72.88% - 82.97%)* | *87.69% (CI: 80.94% - 92.28%)* | *67.46% (CI: 60.07% - 74.06%)* | *84.86% (CI: 81.25% - 88.25%)* | *0.6295* |
| *DeepSeek-R1-Llama70B* | *768 (100.00%)* | *88.61% (CI: 86.02% - 90.78%)* | *93.32% (CI: 90.72% - 95.23%)* | *90.67% (CI: 87.78% - 92.93%)* | *83.33% (CI: 77.42% - 87.94%)* | *77.67% (CI: 71.51% - 82.82%)* | *91.98% (CI: 90.23% - 93.64%)* | *0.7248* |

*Table describes the full document-level analysis results for the various NLP models in the study.*
