## Appendix I for "From Rule-Based to DeepSeek R1 – A Robust Comparative Evaluation of Fifty Years of Natural Language Processing (NLP) Models To Identify Inflammatory Bowel Disease Cohorts"

Appendix I: Full Fairness Results

#### Gender

***Table I-1:*** *Fairness Metrics (Gender) By Model – M vs F*

| *Model* | Demographic Parity (DP) | Equal Opportunity (EO) | Disparate Impact (DI) |
| --- | --- | --- | --- |
| *Regex* | 0.017 | 0.0 | 1.017 |
| *Spacy* | 0.017 | 0.0 | 1.018 |
| *BOW* | 0.064 | 0.015 | 1.080 |
| *TF-IDF* | 0.045 | 0.0 | 1.007 |
| *Word2Vec* | 0.013 | 0.0 | 0.998 |
| *SBERT* | 0.076 | 0.008 | 1.083 |
| *SBERT Med* | 0.053 | 0.009 | 1.058 |
| *DistilBERT* | 0.076 | 0.002 | 1.095 |
| *BioclinicalBERT* | 0.057 | 0.013 | 1.069 |
| *RoBERTa* | 0.0 | 0.0 | 1.0 |
| *Mistral-0.3-7B* | 0.046 | 0.025 | 1.068 |
| *M42-Llama_8B* | 0.053 | 0.034 | 1.077 |
| *DeepSeek-R1- Qwen2.5_32B* | 0.064 | 0.047 | 1.102 |
| *Qwen3_32B* | 0.076 | 0.063 | 1.122 |
| *DeepSeek-R1-Llama70B* | 0.071 | 0.038 | 1.117 |

*Table I-1 describes the full fairness metrics for the gender comparisons in the study.*

***Table***  confirms that the LLMs, particularly Deepseek and Qwen, are the most biased against gender, all favouring men. None of the algorithms breaches the 1.25 threshold, but the larger LLMs breach above DI 1.1.

#### IMD

Significant socioeconomic biases are present and detectable in the LLMs, as per ***Table I‑1*** against the wealthier patients. It is unclear why this happens, but Qwen v2.5 and v3 were particularly biased against the higher sociodemographic groups. These biases are not present in the locally trained models at all. Currently, it is unclear how LLMs are making these biased assumptions about who would be an IBD patient; however, it is clear that such biases are present in the models.

***Table I‑1:*** *Fairness Metrics (IMD) By Model – 1-2 vs 9-10*

| *Model* | Demographic Parity (DP) | Equal Opportunity (EO) | Disparate Impact (DI) |
| --- | --- | --- | --- |
| *Regex* | -0.043 | 0.013 | 0.956 |
| *Spacy* | -0.061 | 0.0 | 0.938 |
| *BOW* | -0.083 | 0.002 | 0.937 |
| *TF-IDF* | -0.002 | 0.0 | 0.992 |
| *Word2Vec* | 0.003 | 0.0 | 1.017 |
| *SBERT* | 0.011 | 0.013 | 0.978 |
| *SBERT Med* | -0.045 | 0.009 | 1.058 |
| *DistilBERT* | -0.047 | 0.026 | 1.031 |
| *BioclinicalBERT* | -0.027 | 0.013 | 0.969 |
| *RoBERTa* | 0.0 | 0.0 | 1.0 |
| *Mistral-0.3-7B* | -0.130 | 0.023 | 0.821 |
| *M42-Llama_8B* | -0.123 | 0.085 | 0.837 |
| *DeepSeek-R1- Qwen2.5_32B* | -0.215 | 0.145 | 0.695 |
| *Qwen3_32B* | -0.234 | 0.169 | 0.670 |
| *DeepSeek-R1-Llama70B* | -0.126 | 0.043 | 0.830 |

*Table I‑1 describes the full fairness metrics for the IMD comparisons in the study.*

#### Age

Substantial age-related biases also appear to be present in only the LLMs who favour the younger patients disproportionately as having IBD per ***Table I-2***. This effect is particularly stark with the larger models who are heavily biased towards younger patients having IBD. However, of note, they did not know the ages of the patients, so they must be attending to something in the text more common in the younger age groups than the older groups. The BERT models and BOW are interestingly biased in the other direction towards the older age groups.

***Table I-2:*** *Fairness Metrics (Age) By Model – 20-30 vs 60-70*

| *Model* | Demographic Parity (DP) | Equal Opportunity (EO) | Disparate Impact (DI) |
| --- | --- | --- | --- |
| *Regex* | -0.014 | 0.0 | 0.985 |
| *Spacy* | 0.011 | 0.0 | 0.994 |
| *BOW* | 0.160 | 0.030 | 1.163 |
| *TF-IDF* | 0.087 | 0.023 | 1.017 |
| *Word2Vec* | 0.019 | 0.0 | 1.010 |
| *SBERT* | 0.101 | 0.030 | 1.123 |
| *SBERT Med* | 0.102 | 0.009 | 1.065 |
| *DistilBERT* | 0.151 | 0.002 | 1.195 |
| *BioclinicalBERT* | 0.168 | 0.030 | 1.214 |
| *RoBERTa* | 0.0 | 0.0 | 1.0 |
| *Mistral-0.3-7B* | -0.030 | 0.133 | 0.958 |
| *M42-Llama_8B* | -0.153 | 0.184 | 0.992 |
| *DeepSeek-R1- Qwen2.5_32B* | -0.097 | 0.268 | 0.859 |
| *Qwen3_32B* | -0.070 | 0.193 | 0.898 |
| *DeepSeek-R1-Llama70B* | 0.010 | 0.253 | 0.756 |

*Table I-2 describes the full fairness metrics for the age-related comparisons in the study.*

#### Ethnicity

The ethnicity results are interesting because they suggest that the larger LLMS are far less biased than, the smaller ones in terms of ethnicity. Unfortunately, the BOW and BERT models are also biased in this regard, likely due to the demographics on which they were trained.

***Table I‑3:*** *Fairness Metrics (Ethnicity) By Model – White vs African*

| *Model* | Demographic Parity (DP) | Equal Opportunity (EO) | Disparate Impact (DI) |
| --- | --- | --- | --- |
| *Regex* | -0.013 | 0.0 | 0.987 |
| *Spacy* | -0.028 | 0.0 | 0.972 |
| *BOW* | 0.039 | 0.177 | 1.248 |
| *TF-IDF* | -0.049 | 0.0 | 1.0 |
| *Word2Vec* | -0.021 | 0.0 | 0.994 |
| *SBERT* | 0.164 | 0.184 | 1.055 |
| *SBERT Med* | -0.059 | 0.010 | 0.941 |
| *DistilBERT* | 0.159 | 0.180 | 1.216 |
| *BioclinicalBERT* | 0.157 | 0.177 | 1.252 |
| *RoBERTa* | 0.0 | 0.0 | 1.0 |
| *Mistral-0.3-7B* | 0.023 | 0.076 | 1.174 |
| *M42-Llama_8B* | 0.125 | 0.052 | 1.215 |
| *DeepSeek-R1- Qwen2.5_32B* | -0.051 | 0.1184 | 0.941 |
| *Qwen3_32B* | -0.041 | 0.114 | 0.926 |
| *DeepSeek-R1-Llama70B* | -0.021 | 0.133 | 0.966 |

*Table I‑3 describes the full fairness metrics for the ethnicity based comparisons in the study.*
