## Appendix F for "From Rule-Based to DeepSeek R1 – A Robust Comparative Evaluation of Fifty Years of Natural Language Processing (NLP) Models To Identify Inflammatory Bowel Disease Cohorts"

Appendix J: Explainability Data

Some of these words map well to the regex and spacy terms such as ‘ulcerative’, ‘crohn’, ‘inflammatory’, ‘azathioprine’, ‘mayo’, ‘crypt’, and ‘vedolizumab’. However, many of them, such as ‘clinic’, ‘allele’, ‘number’, ‘date_time’, ‘with’, ‘normal’, ‘contact’, and ‘caecum’, do not. This suggests that these models have all assigned undue weight to some inappropriate terms. These results are reflected in the top positive feature lists generated from the resulting LR classifiers, which are also quite mixed.

LIME analysis reveals that both distilBERT and bioclinicalBERT weighted terms like ‘UC’, ‘granulomatous’, drug names, and ‘IBD’ positively. However, these models also positively weighted ‘DATE_TIME’ and ‘PERSON’, indicating that the masking tokens from pseudonymisation were likely adversely affecting algorithm performance(84). This might be resolved in future iterations using hide-in-plain-sight pseudonymisation. RoBERTa, by comparison, identified more terms such as ‘kgs’, ‘weekly’, ‘no’, and ‘ongoing’ as positively weighted.

The distilBERT and bioclinicalBERT models make similar mistakes, albeit less dramatically. Both identify the active IBD element but commit different errors. This example highlights how contradictory information within a clinical report can easily confuse a language model.

***Figure J:*** *RoBERTa LIME Error Example*

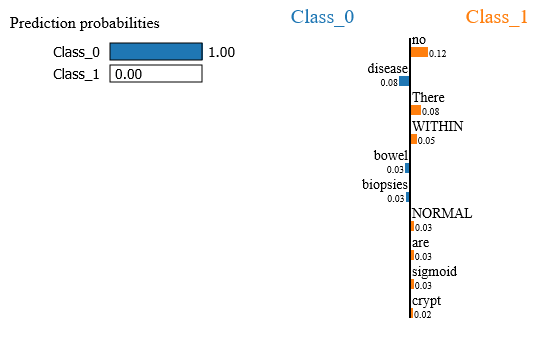

*FigureJ display an informative LIME example of where the RoBERTa model makes some critical mistakes*

The example above is taken from a biopsy sample report in which the clinical history states ?active IBD in a patient with known active IBD; however, the biopsies themselves are normal. A clinician reading this would correctly classify it as IBD in remission (Class 1). However, the model focuses on the incorrect features and classifies the
